# Serological Responses to Human Virome Define Clinical Outcomes of Italian Patients Infected with SARS-CoV-2

**DOI:** 10.1101/2020.09.04.20187088

**Authors:** Limin Wang, Julián Candia, Lichun Ma, Yongmei Zhao, Luisa Imberti, Alessandra Sottini, Kerry Dobbs, NIAID-NCI COVID Consortium, Andrea Lisco, Irini Sereti, Helen C. Su, Luigi D. Notarangelo, Xin Wei Wang

**Affiliations:** Laboratory of Human Carcinogenesis, Center for Cancer Research, National Cancer Institute, Bethesda, Maryland 20892; CCR-SF Bioinformatics Group, Advanced Biomedical and Computational Sciences, Frederick National Laboratory for Cancer Research, 8560 Progress Drive, Frederick, Maryland 21701; CREA Laboratory, Diagnostic Department, ASST Spedali Civili di Brescia, Brescia, Italy; Laboratory of Clinical Immunology and Microbiology, National Institute of Allergy and Infectious Diseases, Bethesda, Maryland 20892; Laboratory of Immunoregulation, National Institute of Allergy and Infectious Diseases, Bethesda, Maryland 20892; Liver Cancer Program, Center for Cancer Research, National Cancer Institute, Bethesda, Maryland 20892

## Abstract

Severe acute respiratory syndrome coronavirus 2 (SARS-CoV-2) is responsible for the pandemic respiratory infectious disease COVID-19. However, clinical manifestations and outcomes differ significantly among COVID-19 patients, ranging from asymptomatic to extremely severe, and it remains unclear what drives these disparities. Here, we studied 159 hospitalized Italian patients with pneumonia from the NIAID-NCI COVID-19 Consortium using a phage-display method to characterize circulating antibodies binding to 93,904 viral peptides encoded by 1,276 strains of human viruses. SARS-CoV-2 infection was associated with a marked increase in individual’s immune memory antibody repertoires linked to trajectories of disease severity from the longitudinal analysis also including anti-spike protein antibodies. By applying a machine-learning-based strategy, we developed a viral exposure signature predictive of COVID-19-related disease severity linked to patient survival. These results provide a basis for understanding the roles of memory B-cell repertoires in COVID-19-related symptoms as well as a predictive tool for monitoring its clinical severity.

## Introduction

Coronavirus disease-2019 (COVID-19) is a severe acute respiratory disease caused by the infection of SARS-CoV-2, which belongs to a group of viruses with a positive-sense, single-stranded RNA genome, similar to the other β-coronaviruses including SARS-CoV (aka SARSCoV-1), MERS-CoV, and other seasonal and less pathogenic coronaviruses (i.e., HKU and OC43) (Gorbalenya et al., 2020; Gussow et al., 2020; Lu et al., 2020; Tian et al., 2020; Wu et al., 2020; Zhou et al., 2020; Zhu et al., 2020). SARS-CoV-2, discovered in January 2020, is directly responsible for more than 25 million confirmed cases and 844,000 deaths globally, as of August 30, 2020. SARS-CoV-2 infection is associated with a wide spectrum of clinical manifestations that range from an asymptomatic infection to a viral syndrome with lower respiratory tract involvement and multifocal interstitial pneumonia, in some cases progressing to severe hypoxic respiratory failure with acute respiratory distress syndrome (ARDS), multiorgan failure and death (Wiersinga et al., 2020). The clinical course and animal models of COVID-19 disease suggest a biphasic trajectory initially dominated by innate and early adaptive immune responses that can result in viral clearance and clinical resolution, but in cases of ineffective early responses, by a persistent and more dysregulated inflammatory response hallmarked by profound morbidity and mortality. It is unclear, however, why some patients are asymptomatic while others have developed severe symptoms. Among hospitalized patients, older patients are more likely to develop severe disease (Goyal et al., 2020; Guan et al., 2020), and patients with cancer are likely to be diagnosed with COVID-19 and have more severe symptoms (Kuderer et al., 2020; Yu et al., 2020; Zhang et al., 2020). As COVID-19-related severe clinical presentations are associated with persistent viral shedding and dysregulated inflammatory response, it is plausible that ineffective host immune responses may contribute to determine the severity of clinical manifestations (Li et al., 2020).

Viruses may affect human health by altering host immunity, contributing to the pathogenesis of inflammatory disorders such as autoimmune disease and cancer (Cadwell, 2015; Foxman and Iwasaki, 2011; Plummer et al., 2016). Various human viruses may interact with one another in the host and may alter a host’s response to new infections and thereby disease severity. It is known that viruses that persist or are cleared in the host may leave unique molecular footprints known as viral epitope-specific reactive antibodies that can affect host susceptibility to other infections, which may be a surrogate of disease severity and progression. For example, prior infection of human herpesvirus 5 (CMV) could improve immune response to influenza (Furman et al., 2015). The recent identification of cross-reactive T cells to SARS-CoV-2 in unexposed individuals may provide a hint about a possible impact of past exposure to seasonal coronaviruses on COVID-19-related outcomes (Braun et al., 2020; Grifoni et al., 2020). Thus, the pattern and type of serological responses to human viruses represent a unique immunological host-specific signature that provides insight into the history of viral exposures, host immunity of each individual, and disease onset.

We have recently employed a synthetic virome technology, VirScan, to detect the exposure history to most known human viruses (Liu et al., 2020). VirScan applies a phage display library that covers 96,179 viral peptides that are each 56 residues in length tilling the protein sequences with 28 residues overlaps, which corresponds to 206 viral species and 1,276 human viral strains (Xu et al., 2015). Using this technology, we developed a viral exposure signature (VES) that could discriminate liver cancer patients from at-risk or healthy individuals and validated the results in a longitudinal cohort with at-risk patients who had long-term follow-up for liver cancer development (Liu et al., 2020). Remarkably, VES could predict cancer among at-risk patients prior to a clinical diagnosis and appeared clinically applicable for liver cancer surveillance. In the present study, we hypothesize that VirScan may provide a sensitive approach to identify immunological footprints predictive of different clinical outcomes of SARS-CoV-2 infection. To that aim, we performed serological profiles of 159 hospitalized patients with pneumonia who were suspected to be infected with SARS-CoV-2, as a part of the NIAID-NCI COVID-19 Consortium study.

## Results

Among 159 hospitalized patients with pneumonia, 156 patients were tested positive while 3 patients were negative for the presence of SARS-CoV-2 infection. In this cohort, 73% of patients were male and the median age was 60 years. Twelve patients were classified as moderate, 19 as severe, and 128 as critical (Table 1). Among the latter, 30 patients died during hospitalization. We used version 2 of the VirScan phage library to profile the Brescia cohort (Fig 1A), which yielded on average 2 million single-end reads per serum sample with mean mapped reads of 91%, comparable to the Maryland (NCI-UMD) cohort recently described (Liu et al., 2020). Using replicates for setting up the reproducibility threshold with built-in blank replica for each plate as background controls, we obtained 16,536 significantly enriched viral epitopes. We detected a median of six species of virus per sample, slightly lower than the median of seven species of virus per sample found among healthy individuals from the Baltimore area in the USA (Liu et al., 2020). Interestingly, while we found a median of 9 viral epitopes per sample in the Brescia cohort, there was an elevation of the total number of unique epitopes across all viruses in COVID-19 patients compared to non-COVID-19 patients, although it did not reach statistical significance (Fig 1B). The prevalence of the most common viruses, such as human respiratory syncytial virus (HRSV), related to respiratory tract infections, human herpesvirus 1 (HHV1), related to oral lesions and encephalitis, human herpesvirus 4 (EBV), related to infectious mononucleosis and human herpesvirus 5 (CMV), was comparable between Brescia COVID-19 patients (Fig 1C) and healthy individuals from Baltimore, as well as other populations (Liu et al., 2020; Xu et al., 2015). Interestingly, we observed some variations of viral compositions among moderate, severe, and critical patients with a tendency of an increasing level of unique viral epitopes corresponding to disease severity (Fig 1C), and this was confirmed by a quantitative analysis of the total amount of viral epitopes across severity patient groups (Fig 1D-E). There was a statistically significant increase in reactive viral epitopes with increasing severity in clinical presentations. Noticeably, SARS-CoV was detected in 27% of moderate patients but 53% of severe patients and 49% of critical patients (Fig 1C, Suppl Table S1). The difference of viral composition and levels of reactive unique epitopes was comparatively small when stratified by age and sex (Suppl Fig S1). Taken together, these results indicate that an individual’s immune memory antibody repertoires due to a history of viral exposure showed a marked increase during acute SARS-CoV-2 infection.

**Table 1.** Clinical Characteristics of Hospitalized Patients with Pneumonia from Brescia.

| Variable | All | Moderate | Severe | Critical |
| --- | --- | --- | --- | --- |
| SARS-CoV-2 status<sup>1</sup> - no. (%) |  |  |  |  |
| Negative | 3 (2) | 1 (33) | 0 | 2 (66) |
| Positive | 156 (98) | 11 (7) | 19 (12) | 126 (81) |
| Sex - no. (%) |  |  |  |  |
| Male | 116 (73) | 7 (58) | 8 (42) | 101 (79) |
| Female | 43 (27) | 5 (42) | 11 (58) | 27 (21) |
| Age - median (range) | 60 (23-89) | 56 (32-89) | 57 (24-84) | 60.5 (23-83) |
| Sample time point<sup>2</sup> - no. |  |  |  |  |
| T0 | 159 | 12 | 19 | 128 |
| T1 | 76 | 1 | 4 | 71 |
| T2 | 41 | 0 | 2 | 39 |
| T3 | 22 | 0 | 0 | 22 |
| Outcome - no. (%) |  |  |  |  |
| alive | 129 (81) | 12 (100) | 19 (100) | 98 (77) |
| deceased | 30 (19) | 0 | 0 | 30 (23) |

<sup>1</sup> SARS-CoV-2 status was determined by quantitative real-time reverse-transcriptase-polymerase-chain-reaction (qRT-PCR) assay of nasal and pharyngeal swab specimens and/or by positive serology tests.
<sup>2</sup> Longitudinal time points of blood collection are indicated as T0 - T3. T0, the first blood collection; T3, the last blood collection.

**Figure 1.**
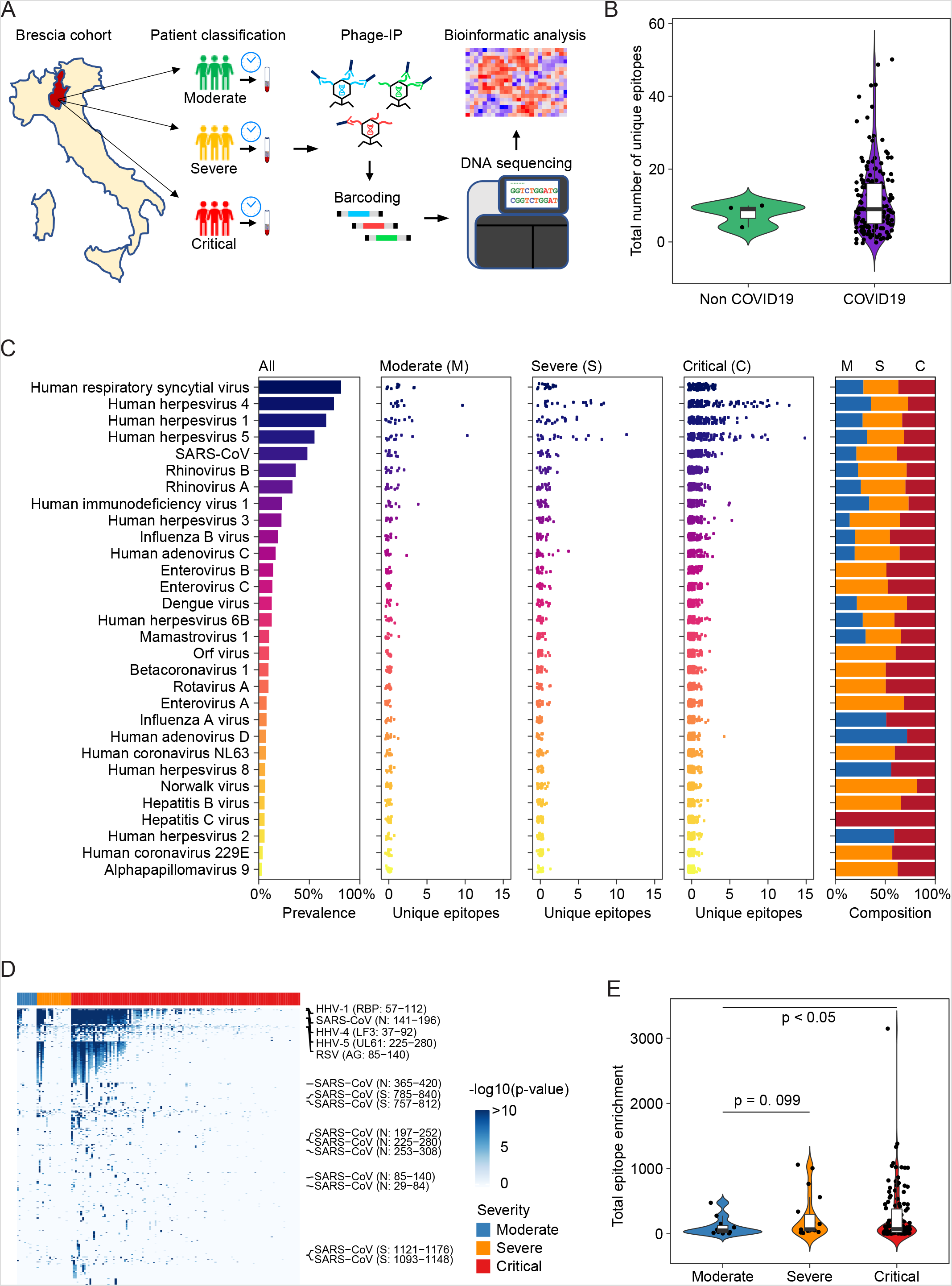
Overview of viral exposure of the Brescia cohort. (A) Workflow of VirScan. (B) The total number of unique epitopes in non-COVID and COVID-19 cases. (C) Prevalence of viruses of 156 COVID-19 cases (left), the number of unique epitopes of moderate, severe and critical groups of patients (middle 3 panels) as well as the composition of prevalence of the three groups of patients (right). (D) Antibody reactivity of all the epitopes presented in at least two cases. Each row represents an epitope while each column stands for one CODIV-19 case. –log10 (pvalue) was used to measure the enrichment of peptide. (E) The total enrichment of all epitopes in the moderate, severe, and critical groups. Log transformation was applied. In violin plots, boxes span the interquartile range; lines within boxes represent the median; the width of violin plots indicates the kernel density of values.

Nearly 50% of COVID-19 patients from Brescia had antibodies reactive to SARS-CoV peptides (Fig 1C, left panel). We also detected antibodies reactive to peptides corresponding to several other members of coronavirus including NL63, 229E, and HKU1, but their prevalence was low (7% NL63, 4% 229E, 2% HKU1). The levels of SARS-CoV positive cases were unexpectedly high in Brescia patients, given the fact that reactive antibodies to SARS-CoV epitope sequences were hardly detectable in our previous VirScan study of 899 Baltimore patients and other populations, suggesting that the SARS-CoV prevalence is extremely low in the general population (Liu et al., 2020; Xu et al., 2015). Consistently, SARS-CoV reactivity could not be detected in three Brescia patients diagnosed with pneumonia but tested negative for SARS-CoV-2 while its level was high in COVID-19 patients (Fig 2A). We hypothesized that the SARS-CoV peptides used in the Phage libray may detect SARS-CoV-2 instead of SARS-CoV. We examined closely the VirScan phage library and found that it contained 80 epitope tiles corresponding to the SARS-CoV sequence, namely 44 epitopes for the spike protein, 15 epitopes for the nucleocapsid protein, and 21 epitopes for 3a, 3b, 7a, and 9b proteins. Among them, four peptides spanning the spike protein sequence and 8 peptides spanning the nucleocapsid protein showed strong reactive signals to COVID-19 positive patients (Fig 2B). Since SARS-CoV and SARSCoV-2 share a similar viral structure (Fig 2C) with an overall 79% genetic similarity, we determined sequence similarities between SARS-CoV epitopes used in the library and the newly identified SARS-CoV-2 sequences. We found that reactive SARS-CoV epitope sequences share a high homology to SARS-CoV-2, especially in the regions predicted to be strongly antigenic by the B-cell epitope prediction tool of the Immune Epitope Database (http://tools.iedb.org) (Fig 2C-F). Thus, the VirScan’s SARS-CoV peptides may be effectively used to detect SARS-CoV-2 antibodies. We measured the antibody epitope binding signal (EBS) to estimate antibody titer for each epitope recently described (Mina et al., 2019). Similar to an overall increase in the immune memory antibody repertoire diversity to all known pathogenic viruses corresponding to clinical severity, we found a significant increase in levels of antibodies to the spike protein, but not to the nucleocapsid protein (Fig 2G). We also found an increasing trend in antibody production to the spike protein over time during hospitalization (Fig 2H). Consistently, we found a significant increase in antibodies to the spike protein in patients requiring intensive care unit (ICU) care compared with non-ICU patients (Suppl Fig S2A). However, no statistical difference was observed for antibodies against the nucleocapsid protein. Moreover, levels of anti-S antibodies did not correlate with mortality (Suppl Fig S2B-C).

**Figure 2.**
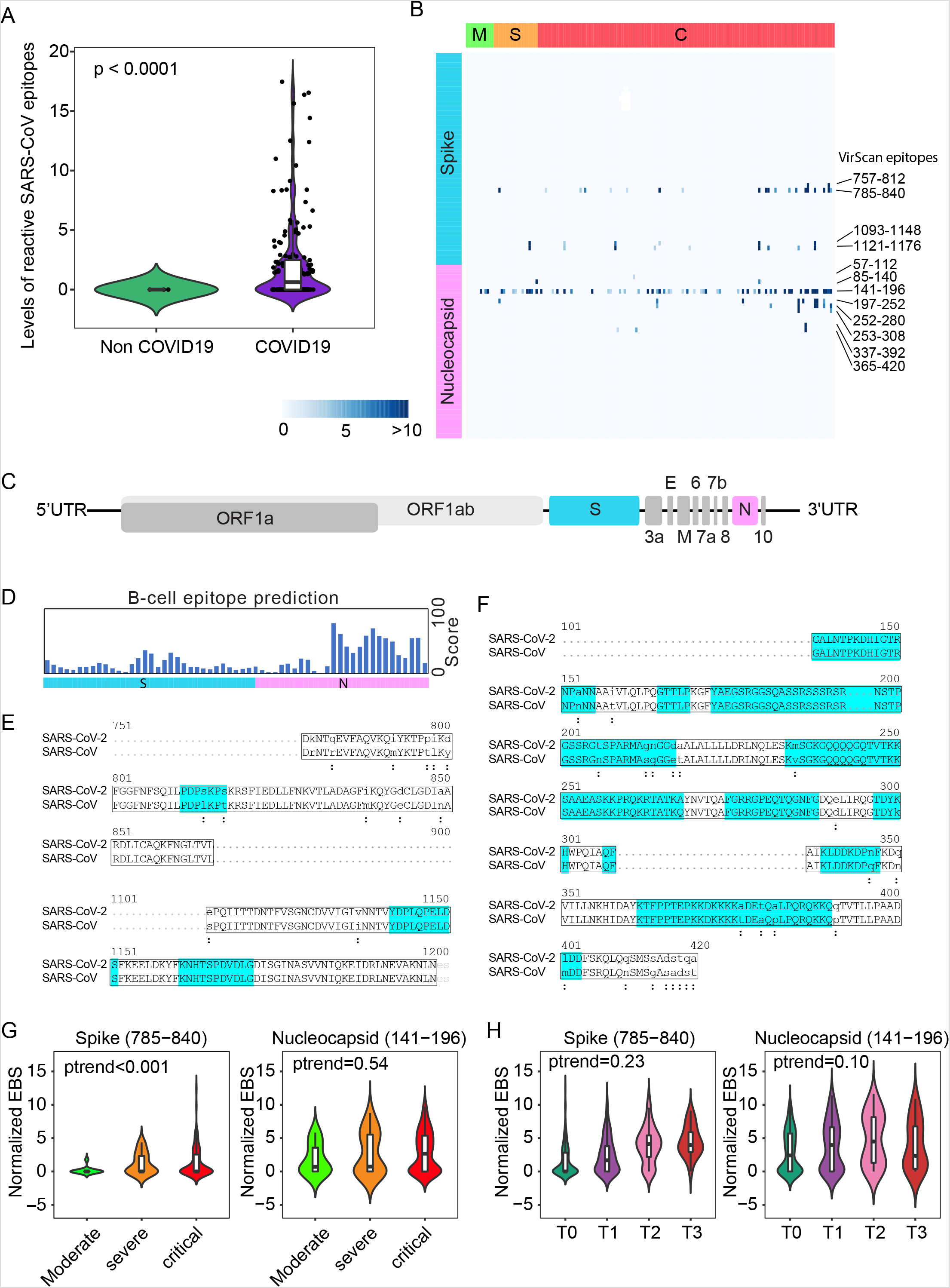
SARS-CoV epitope reactivity in moderate, severe, and critical cases. (A) Total reactivity of all SARS-CoV epitopes in non-COVID and COVID-19 cases. Log transformation was applied. (B) Antibody reactivity of 85 SARS-CoV epitopes. Each row represents the significant peptide tiling corresponding to spike protein (1–1255) and nucleocapsid protein (1–422). The color intensity of each cell corresponds to the scaled –log10(p value) measure of significance of enrichment for a peptide in a sample. (C) Organization of SARS-CoV-2 genome encoding various viral proteins. (D) B-cell epitope prediction score for spike and nucleocapsid based on the Immune Epitope Database and Analysis Resource (IEDB). (E-F) Sequence alignment of reactive peptides corresponding to spike (E) and nucleocapsid protein (F) of SARSCoV and SARS-CoV-2. Only peptide sequences in the phage library are shown. Residues with perfect match are capitalized. Predicted epitopes by IEDB are highlighted. (G) Normalized EBS of spike (785–840, left) and nucleocapsid (141–196 right) proteins in moderate, severe and critical patients. (H) Normalized EBS of spike (785–840, left) and nucleocapsid (141–196 right) proteins in patients at different time points. In violin plots, boxes span the interquartile range; lines within boxes represent the median; the width of violin plots indicates the kernel density of values.

The above results suggest that an individual’s immune memory antibody repertoires may be modulated upon acute infection by SARS-CoV-2. To further explore this hypothesis, we tracked the longitudinal progression of EBS for individual patients. Fig 3A shows the individual trajectories over time for COVID-19 patients grouped by disease severity (gray lines), which were generated from discrete timepoints using LOESS regression. The average trajectory for each group (solid blue line) was then fitted by linear regression (dashed blue line) to extract the slope’s value and standard error (shown in the legend). While the rate of change of the moderate and severe groups is almost flat (i.e. the slope is consistent with zero after taking the standard error into account), the critical group shows a significant positive slope. Therefore, in addition to having an increased EBS at baseline, COVID-19 patients in the critical group display a characteristic overall increase across the entire antibody repertoire over time. Consistent with these observations, the EBS rate of change among ICU patients is observed to be significantly larger compared with that of non-ICU patients (Suppl Fig S3A) and, analogously, the EBS slope among deceased patients appears significantly larger than that of survivors (Suppl Fig S3B). Furthermore, similar analyses of sex and age effects on the humoral immune response of COVID-19 patients show that both the total epitope enrichment at baseline (Suppl Fig S4A-B) and the longitudinal EBS progression (Suppl Fig S4C-F) increase with age and are significantly higher among males compared with female patients. In agreement with well-established observations of older patients, preferentially male, being more susceptible to severe COVID-19 progression and death (Goyal et al., 2020; Guan et al., 2020; Takahashi et al., 2020), our findings solidify the emerging picture of disease severity being robustly associated with an elevated, nonspecific humoral immune activation. Fig 3B shows, from left to right, the correlation of the EBS signal calculated across all epitopes (horizontal axis) against the EBS signal from the 785–840 epitope in the spike protein, the 141–196 epitope in the nucleocapsid protein, and all available SARS-CoV peptide sequences, respectively. As expected, restricting the EBS signal to one or a few epitopes is noisier, but nonetheless we observe significant positive correlations with the overall EBS across the cohort. The heatmap in Fig 3C shows in greater detail the integrated longitudinal EBS progression from the three patient groups (indicated by the color bar on the left), which emphasizes the intriguing complexity and heterogeneity of the COVID-19 time course.

**Figure 3.**
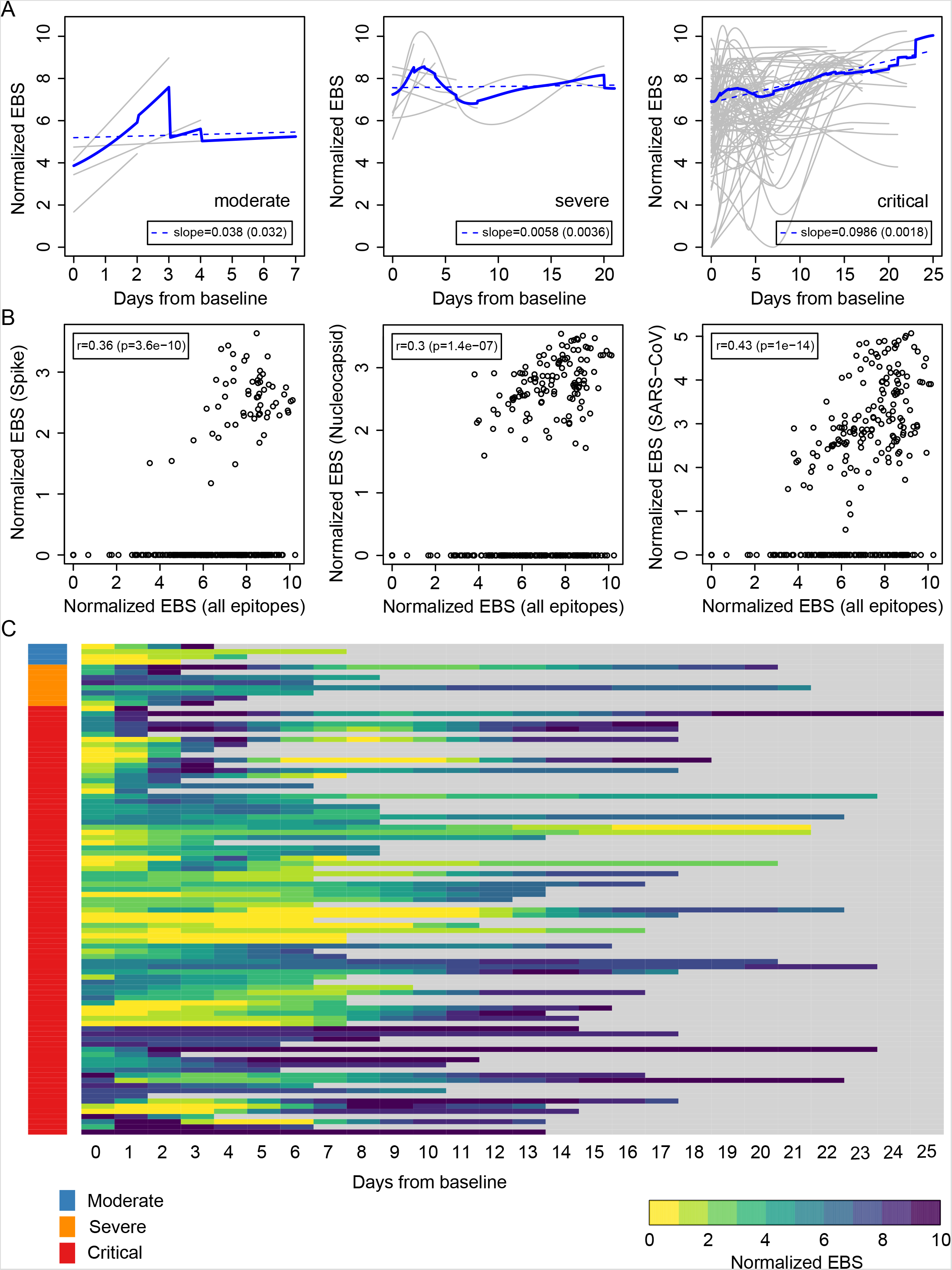
Longitudinal progression of the normalized EBS across individuals. (A) Individual trajectories over time for patients grouped by disease severity (gray lines), which were averaged (solid blue line) and fitted by linear regression (dashed blue line; slope and standard error shown in the legend). Baseline refers to the first sample obtained after admission to the hospital. (B) Normalized EBS in SARS-CoV spike, nucleocapsid, and all SARS-CoV reactive proteins compared against the normalized EBS across all VirScan epitopes. (C) Heatmap showing the longitudinal progression of individual patients integrated across all three patient groups by disease severity.

To determine if virus-related memory antibody repertories could be used to define COVID-19 clinical manifestations predictive of clinical outcomes, we utilized a gradient boosting machine learning approach, XGBoost, to build a COVID-19-related VES (COVID-VES) corresponding to the severity of patients’ conditions, a strategy as before (Liu et al., 2020). We only included baseline samples (i.e., the first sample available upon hospitalization) to search for COVID-VES in order to minimize the potential disturbance of serological responses due to acute infection by SARS-CoV-2 or due to the length of hospitalization. Patients with a moderate to severe condition were compared to those with a critical condition. A total of 100 iterations were performed by applying the algorithm ROSE to generate balanced classes; for each iteration, XGBoost with 10-fold cross validation was found capable to significantly discriminate moderate samples from critical samples with area-under-the-curve (AUC) performance values close to 1 during training and above 0.9 during cross-validation (Fig 4A). The resulting COVID-VES signature consisted of 28 viruses that were selected in at least 50 of the 100 iterations. All of these viral strains were positively associated with patients with critical condition (Fig 4B). We then tested if COVID-VES is associated with overall survival by applying the survival risk prediction algorithm that was successfully used in previous studies (Ma et al., 2019; Roessler et al., 2010). The survival risk prediction based on 10-fold cross-validation predicted patients into low- and high-risk groups with a significant difference in survival, as shown in the Kaplan-Meier plot (Fig 4C), resulting in the log-rank P-value = 0.008. The cross-validated misclassification rates were significantly lower than expected by chance (permutation P-value = 0.04 based on 100 random permutations). The Cox proportional hazards regression analysis was stratified by several clinical groups. We found that ICU, age older than 60 years, and male sex were associated with overall survival (Fig 4D). We did not perform multivariable Cox regression analysis due to the small size of our cohort.

**Figure 4.**
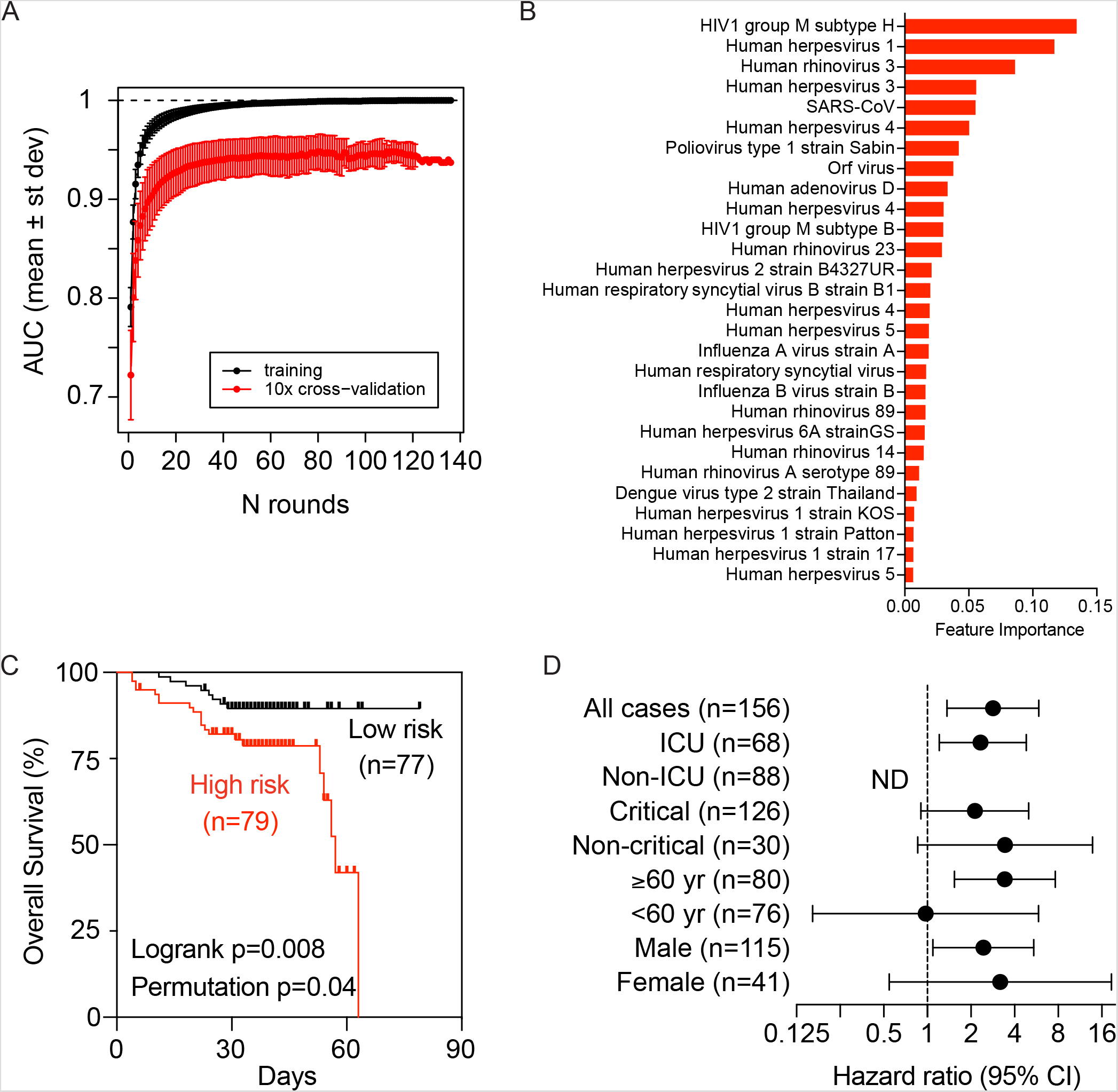
The development of a viral exposure signature predictive of disease severity. (A) XGboost with 10-fold cross validation for 100 iterations of balanced input data generated by ROSE. Each iteration showing the AUC value of training and cross validation sets. (B) COVIDVES signature consisted of 28 viruses that were selected in at least 50 of the 100 iterations predicted by XGboost. (C) Survival risk prediction based on COVID-VES signature viruses in low and high-risk patient groups. Survival time was based on days since admission. (D) The results from Cox proportional hazards regression analysis. ND, not determined.

## Discussion

Cellular immunity plays an important role in antiviral response by producing antibodies against various pathogens such as SARS-CoV-2, among others, which may result in convalescence. Using VirScan, we have determined the exposure history of COVID-19 patients to most known human viruses. Our findings suggest that the dysregulation or imbalanced activation of cellular immunity may be associated with poor COVID-19 outcomes. A surprising finding of this study is a marked increase in the overall immune memory antibody repertoire activity in COVID-19 patients linked to the trajectories of disease severity. This conclusion is supported by the following observations. First, levels of total reactive antibodies against unique epitopes of known viruses were much higher in hospitalized COVID-19 patients than non-COVID-19 patients who were also hospitalized due to pneumonia. Second, COVID-19 patients with critical condition had much higher levels of reactive antibodies to known viruses than those with severe or moderate conditions. These results suggest that COVID-19 patients in critical condition may have a uniquely different host immune response and presumably a different host genetic background. This view is consistent with a recent genomewide association study identifying a 3p21.31 gene cluster as a genetic susceptibility locus in patients with COVID-19 with respiratory failure (Ellinghaus et al., 2020) and with the recent identification of monogenic defects of type I interferon immunity in patients with critical COVID-19 (Zhang et al., submitted). Third, longitudinal analysis revealed that during their hospitalization, patients in critical condition showed the highest elevation of reactive antibodies to known viruses compared to patients in severe or moderate condition. It appears that the elevated immune memory antibody repertoire activity is associated with poor clinical trajectories in COVID-19 patients. We also found that levels of antibodies against an epitope corresponding to SARS-CoV spike protein with 100% homology to SARS-CoV-2 were also linked to trajectories of disease severity. There was no significant difference in immune memory antibody repertoires between older or younger patients but a small difference between men and women who had acute infections of SARS-CoV-2. This is in contrast to the observations that both male and older individuals are more likely to experience severe COVID-19-related symptoms than female or younger individuals (Scully et al., 2020). Our data suggest that SARS-CoV-2 may directly stimulate an individual’s overall immune memory antibody repertoire activity. These results are unexpected since humoral immunity is thought to be very stable over time due to long-lived plasma cells since long-term antibody responses are critical for protective immunity against pathogens (Amanna et al., 2007; Halliley et al., 2015; Inoue et al., 2018). Interestingly, a recent study revealed that measles virus infection can diminish preexisting antibodies that offer protection from other pathogens, which may in turn create potential vulnerability to future infections (Mina et al., 2019). It seems that COVID-19 acute infection and measles virus infection may follow different molecular mechanisms to alter humoral immune memory. Consistently, a recent study demonstrated that COVID-19 infection is associated with increased frequencies of proliferation of memory B cell subsets but no changes in naïve B-cell frequencies (Mathew et al., 2020). The ability of SARSCoV-2 to selectively promote proliferation of memory B cells could explain our findings of a marked increase in an overall immune memory antibody repertoire activity linked to poor clinical trajectories of COVID-19 patients. Future studies will explore whether dampening COVID-19-induced reactivation of memory B cells may be a viable strategy to control the clinical severity of COVID-19 patients. Encouraging results were obtained with the use of dexamethasone (Group et al., 2020), a synthetic corticosteroid as a broad-spectrum immunosuppressor that can affect both T cells and B cells. Inhibition of Bruton tyrosine kinase, a regulator of B-cell maturation (Rip et al., 2018), could be another viable therapeutic strategy to improve clinical outcomes of COVID-19 patients (Roschewski et al., 2020).

Autoimmune diseases are complex disorders resulting from the failure of immunologic tolerance leading to an immune response against the host antigens. Autoimmune reactions signify an imbalance between effector and regulatory immune responses (Rosenblum et al., 2015). They may arise from a combination of genetic and environmental factors. Viral infection is considered as an environmental factor to trigger autoimmune disease (Munz et al., 2009). Examples of human viruses include HHV-4 (EBV) and HHV-6 linked to multiple sclerosis (Soldan et al., 1997; Thacker et al., 2006), parvovirus B19 linked to rheumatoid arthritis (Takahashi et al., 1998), and hepatitis C virus linked to cryoglobulinaemia (Roccatello et al., 2018). Several recent studies suggest a possible involvement of COVID-19 in autoimmune and autoinflammatory diseases such as Kawasaki-like disease in children infected with SARS-CoV-2 (Ehrenfeld et al., 2020; Galeotti and Bayry, 2020). It is plausible that various clinical manifestations of COVID-19 may be the result of SARS-CoV-2-induced autoimmunity. While how COVID-19 induces autoimmunity remains unclear, molecular mimicry has been suggested as a plausible mechanism (Munz et al., 2009). This mechanism suggests the presence of similar antigens between viruses and hosts to facilitate pathogens to avoid the host immune response and is mainly mediated via a T-cell response (Munz et al., 2009). However, our results suggest that COVID-19 may affect B-cell repertoires as we observed a marked elevation of memory antibodies to past history of viral infection of most known viruses, regardless of viral types. These results are consistent with the idea that SARS-CoV-2 may directly activate memory B cells, a concept supported by a recent observation that COVID-19 infection is mainly associated with increased frequencies of proliferation of memory B-cell subsets and expansion of plasmablasts (Mathew et al., 2020). It is possible that memory B cells are much more sensitive than naïve B cells to SARS-CoV-2 and that patients with activation of certain memory B cell subsets may be more vulnerable for COVID-19-induced disease severity. This could explain our finding that HCV infection was seen exclusively in hospitalized Brescia COVID-19 patients with a critical condition. It will be interesting in determining if HCV patients who have achieved a sustained virologic response by anti-HCV therapy are still vulnerable for COVID-19-mediated disease severity. Additional prospective studies with larger cohorts are warranted to test these hypotheses.

In summary, by determining serological responses to history of infection from most known human viruses, we linked the activity of memory antibody immune repertoires to clinical manifestations of COVID-19 patients. We developed a COVID-19-related viral exposure signature as serological biomarkers that may be useful to identify COVID-19 patients who may progress to autoimmune and autoinflammatory disease that may require tailored treatments. The main limitation of this study is the inclusion of only 156 COVID-19 patients without asymptomatic individuals or patients with mild symptoms infected with SARS-CoV-2. While we observed a consistent and clear elevation of individual’s immune memory antibody repertoires linked to trajectories of disease severity over time, there remains a substantial heterogeneity within each patient group, which will require expanding our analysis in the future to larger cohorts with increased time resolution in order to gain a deeper understanding of the complex interplay between host immunity and SARS-CoV-2 infection.

## Data Availability

Data will be made available upon request

## Acknowledgements

We are grateful to all study participants, as well as clinicians, nurses, and study coordinators who helped with patient enrollment during COVID-19 pandemic. We thank Bao Tran, Jyoti Shetty and their team for sequencing support. We also thank Stephen Elledge for providing the VirScan phage library. This work was supported in part by grants from the Intramural Research Program of the Center for Cancer Research of the National Cancer Institute (grant ZIA BC 011951 to XWW), the Division of Intramural Research of the National Institute of Allergy and Infectious Diseases (grant AI001270–01 to LDN) and by Regione Lombardia, Italy (project “Risposta immune in pazienti con COVID-19 e comorbiditá” to LI).

## Author Contributions

LW and XWW developed the study concept. LI and AS contributed patient samples and deidentified metadata. LW performed phage-IP experiments. LW, JC, LM, YZ and XWW performed bioinformatics analysis and data interpretation. KD, AL, IS, HCS, and LDN performed additional data analysis and provided overall clinical assessment. XWW wrote the manuscript with the help from LW, JC, LM and YZ. JC coordinated the study in the framework of the NIAID-NCI COVID-19 consortium. Members of the NIAID-NCI COVID Consortium helped with the organization of the Consortium activities and assessment of clinical metadata. All authors read, edited and approved the manuscript.

## Materials and Methods

### Brescia Cohort Patient Samples

Sera were prepared from blood donated by patients admitted at Spedali Civili in Brescia, Italy due to suspected COVID-19 since March 2020. Deidentified discarded blood samples and clinical classification of patients into categories of mild, moderate, severe, and critical according to clinical features (NationalHealthCommission&NationalAdministrationofTraditionalChineseMedicine, 2020), were obtained under protocol NP-4000 approved by Comitato Etico Provinciale, Brescia, Italy. Patient clinical information and eligibility were surveyed with the standard COVID-19 Human Genetic Effort Patient Screen form. Sera were isolated from whole blood for analysis.

### T7 Phage Epitope Library

The sera samples from enrolled patients were used to perform high through-put phage-immunoprecipitation (Phage-IP; VirScan) with a T7 phage library expressing epitopes of humoral virome. We used the v2.0 epitope library which consist of 93,904 viral peptides generated from virome of all 1,276 known humoral virus strains^17^. In this phage library, there are 3,663 epitopes from 16 species of Corona virus, including 80 epitopes from SARS-CoV. The virome was tiled to 56 amino acids length epitopes with 28 amino acids overlapping. The epitope tiling was then cloned to a T7 phage library for displaying. Phage library was amplified using BLT5403 E. coli strain with plate lysate method as previously described^18^. The phage library quality was assessed by two ways. First, it is tittered with standard protocol to ensure the titer of phage is about 1×10^11^pfu/mL. Second, a 10µl aliquot of the amplified phage library was lysed by boiling at 95°C for 10 min followed by two steps of PCR to amplify and index the cloned epitope sequences in the phage genome. The constructed sequencing library was then sequenced at the NCI CCR Frederick Sequencing Facility. The phage library passed quality control as a total coverage of more than 99.99% of the designed epitopes was achieved. These sequencing results also serve as input for downstream analysis of Phage-IP-seq.

### Phage-IP-seq

Phage-IP was then performed using collected sera samples and phage library as previously described^17^. The day before Phage-IP, 96-deep-well plates (BradTech, Catalog #EW-07904–04) were blocked using bovine serum albumin in TBST buffer. After overnight incubation, the blocking buffer was aspirated and sera samples containing 2 µg of total IgG were mixed with 2×10^10^ pfu T7 library in 1mL dilution buffer (20 mM Tris-HCl, pH 8.0, 100 mM NaCl, 6 mM MgSO4) supplemented with 50 µg/mL chloramphenicol and 50µg/mL kanamycin into the deep well plate, and rotated at 4°C for 20 hours for the phage to form complex with the antibodies from sera. Technical duplicate plates were introduced during this step. After overnight complex formation, 20 µl protein A and 20 µl protein G Dynabeads (Thermo Fisher, Catalog #10008D and #10009D) were added to each well containing the phage and antibody complex, and rotated for another 4 hours at 4°C. The finally formed Dynabeads-antibody-phage complex was then washed three times with washing buffer (50 mM Tris-HCl, pH7.5, 150 mM NaCl, 0.1% NP-40) to eliminate the non-specific binding. After wash, the Dynabeads with antibody-phage complex were suspended in 40 µl ultrapure water and transferred into a 96-well PCR plate, followed by boiling at 95°C for 10min to recover the phage genomic DNA. The recovered phage genomic DNA was used as template of first round PCR to amplify the epitope expressing sequences. A second round PCR was employed to add index barcode to the DNA product of first round PCR of each sample well on the 96-well plate. The products of second PCR were then pooled together and loaded to a 2% agarose gel and target size fragment were cut and recovered using a gel extraction kit (Qiagen, Catalog #28704). With this, the DNA sequencing library was successfully constructed. Recovered sequencing library now contained barcoded DNA fragments encoding viral epitopes that were recognized by antibodies in the sera. This DNA library was then sequenced the same way as phage epitope library did, on an Illumina NextSeq 500 platform with 75bp single read, at NCI CCR Frederick Sequencing Core Facility. A total of 200 million reads per lane were obtained with an average of one million reads per serum sample and minimum 90% mapping rates were achieved.

### Sequencing Data Processing and Informatic Epitope Score Analysis

Raw data were demultiplexed with BCL2FASTQ2 and converted to fastq format. The fastq files were mapped to the virome library sequence with Bowtie. To call hits, we first calculated P-value of each epitope by fitting the observed post-alignment read count abundance distribution of phage-IP enriched epitopes in to a zero-inflated Generalized Poisson null distribution regression model. Then initial QC was performed by using scatterplots of the –log_10_(P-values) and a sliding window width of 0.005 from 0 to 2 across the axis of one replicate. A threshold of –log_10_(P-value) was set between 2.3 and 4.7 in both replicates based on –log_10_(P-value) distribution to correct batch effect and background noises. A hit was called to reference epitope if present in both technical replicate samples. Hits that were present in less than two sera samples or more than three mock immunoprecipitation wells were eliminated as background noise. The called epitope hits were then grouped to the virus it is derived. Another threshold was also used to bioinformatically remove cross-reactive antibodies: called viruses were sorted by total hits number in descending order, and then iterate through each virus in this order to remove any called epitope that shares more than seven amino acids homology with epitopes of previous virus in this list. All left epitopes and virus are now used as specific signal of each sample. The number of epitope hits of each virus after phage-IP are summed and used as the so-called feature score of each virus. As a complementary epitope binding quantitation strategy and to overcome possible limitations of the feature score calculation, we also employed a recently developed strategy to calculate z-scores, by comparing antibody-enriched libraries to replicate negative controls (mock IPs). By determining the observed abundance excess relative to the background, divided by the background signal’s standard deviation, and then converting to log scale, we assessed the EBS.

### Epitope Abundance Count and Virus Prevalence

Epitopes with a feature score greater than one were counted as unique epitopes. Any virus with more than one non-cross-reactive epitope detected is counted as positive. Detected viruses are then counted at strain level according to epitopes detected and grouped to species level. Unique epitopes and grouped detected virus species are then counted in each sample to plot the unique epitope abundance and virus prevalence. Number of detected SARS-CoV epitope are extracted and used for heatmap plot and subsequent analysis.

### B-cell epitope antigenicity prediction

Sequences of SARS-CoV spike and nucleocapsid proteins were used for prediction of B-cell antigenicity with the B-cell epitope prediction tool of the Immune Epitope Database (http://tools.iedb.org). A B-cell antigenicity score of 1–100 for each epitope was predicted by the online software.

### Sequence alignment

Protein sequences of SARS-CoV and SARS-CoV-2 are aligned with the EBI Clustal Omega Multiple Sequence Alignment tool (https://www.ebi.ac.uk/Tools/msa/clustalo/) and NCBI BLASTP tool (https://blast.ncbi.nlm.nih.gov/). Cosmetic labeling was made to highlight the region of interest in the sequences.

### Quantification and Statistical Analysis

Statistical differences and significances between patient groups of moderate, severe and critical are examined with Xgboost algorithm and survival analyses were done with GraphPad Prism 8. P-values were calculated with Student’s t-test or log rank t-test. All plot figures were generated with GraphPad Prism 8, R3.5.3 or R3.5.1 based BRB-ArrayTools Stable Version 4.6.1.

### Analysis of Longitudinal EBS Progression

The normalized EBS signal across all epitopes was analyzed longitudinally for all individuals with two or more timepoints. For those with two timepoints, the normalized EBS signal was linearly interpolated. For those with more than two timepoints, data were fitted using LOESS (Locally Estimated Scatterplot Smoothing) regression. The fitted curves were then averaged within patient groups. Linear regression was performed on the averaged profiles to extract the slope’s value and standard error.

### XGBoost Regression of Viral Exposure Signature

To find out possible viral exposure signature that is associated with severity of COVID-19 patients, we employed open source software named XGBoost, which implements machine-learning algorithms under parallel gradient tree boosting framework^18^. To prepare our data for XGBoost modeling, we first needed to overcome the imbalance problem of our dataset. In our Brescia cohort, there are 11 moderate cases, and 126 critical cases. This would largely impair the performance of classification. Overcoming this imbalance problem, we employed the well-known Random Over-Sampling Examples (ROSE) R package (N. Lunardon, G. Menardi, and N. Torelli. R package ROSE: Random Over-Sampling Examples (version 0.0–3). Università di Trieste and Università di Padova, Italia, 2013. URL http://cran.r-project.org/web/packages/ROSE/index.html.). The ROSE package provides functions to deal with binary classification problems in the presence of imbalanced classes by generating artificially balanced samples according to a smoothed bootstrap approach and allow for aiding both the phases of estimation and accuracy evaluation of a binary classifier in the presence of a rare class. Since ROSE package is very good at dealing with binary dataset, we used the epitope binding score VirScan data of moderate and critical groups. We successfully generated well balanced datasets consisting of moderate (N = 70) and critical (N = 70) groups by 100 iteration. This 100 ROSE balanced datasets were then used for XGBoost regression. We used grid search strategy in XGBoost to maximize the computed mean AUC value. The AUC was generated with 10-fold cross validation where 90% of samples were used for training and the remaining 10% of samples were used as independent validation. To avoid over-fitting, we set the early stop of model training to at least 20 rounds when no incremental improving was observed in the AUC. With this optimized iteration model, XGBoost conducted feature selection and output viral exposure signatures consisting of 56 viruses which could discriminate the critical versus the moderate group of COVID-19 patients in total of 100 iterations. Among the 56 viruses, 28 were the common viruses that were predicted at least 50% of the 100 iterations.

### VES Based Survival Prediction

We used the 28-shared virus exposure signature referred to as COVID-VES to predict survival of Brescia cohort. The survival risk prediction tool of the BRB-ArrayTools Stable Version 4.6.1 was used for survival analysis. Two risk groups at median cut off were selected with 10-fold cross validation method and 100 permutations were performed. With a permutation P-value 0.03, the output survival risk prediction of the 156 training samples were then used for survival analysis for different subgroups of the cohorts with GraphPad Prism 8. Hazard ratio and 95% confidential interval was used for forest plot of the different groups.

### Additional Statistical Methods

All other analysis was performed with GraphPad Prism 8 or R 3.6.3. Comparison of different groups were analyzed with two-sided Student’s t-test.

## Supplementary Figure Legends

**Figure S1.**
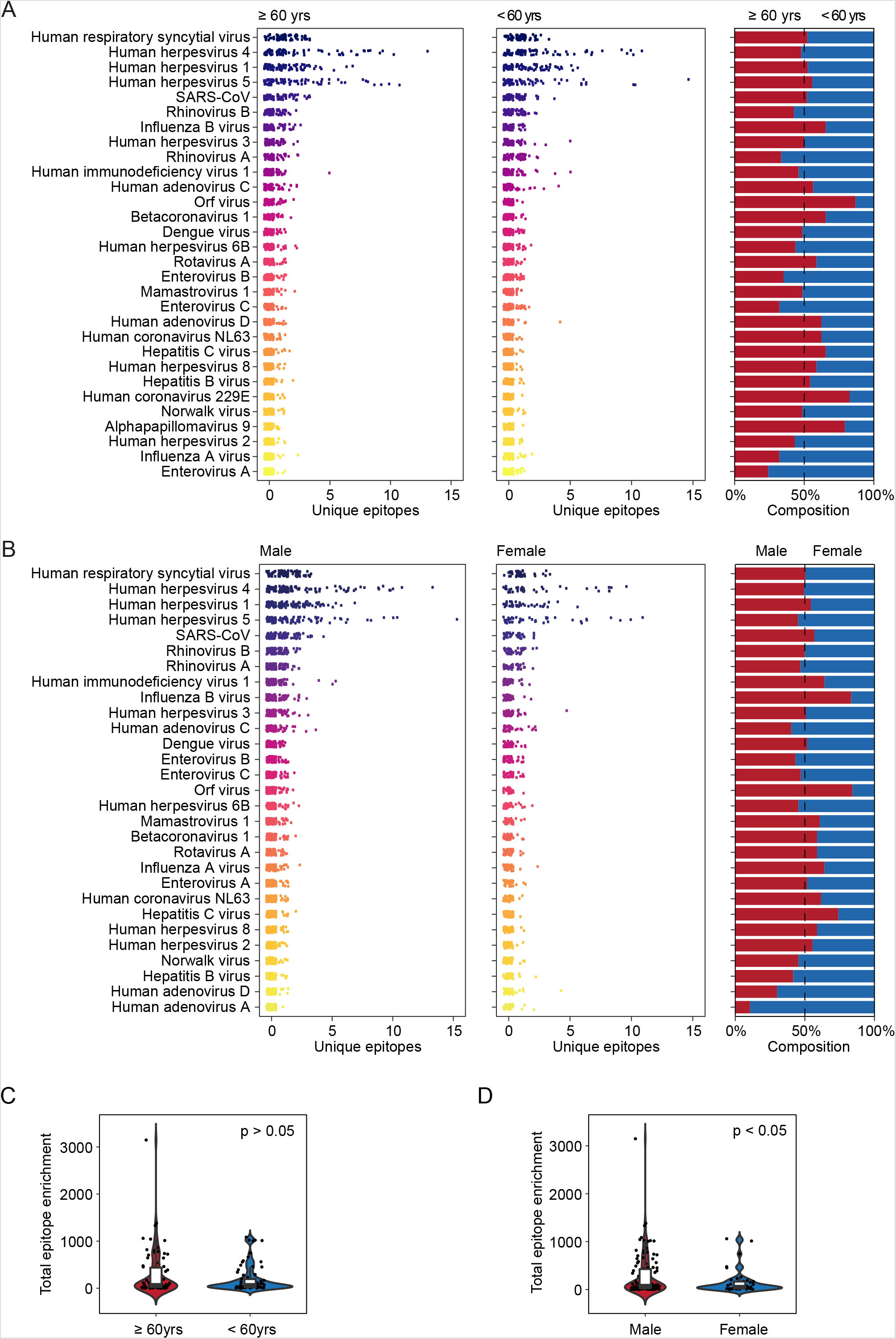
Prevalence of viruses. (A) The number of unique epitopes and the composition of prevalence in cases with age ≥ 60 yrs (the median of all COVID cases) and age < 60 yrs. (B) The number of unique epitopes and the composition of prevalence in male and female cases. (C) The total reactivity of all epitopes in cases with age ≥ 60 yrs and age < 60 yrs. (D) The total reactivity of all epitopes in male and female cases. In violin plots, boxes span the interquartile range; lines within boxes represent the median; the width of violin plots indicates the kernel density of values.

**Figure S2.**
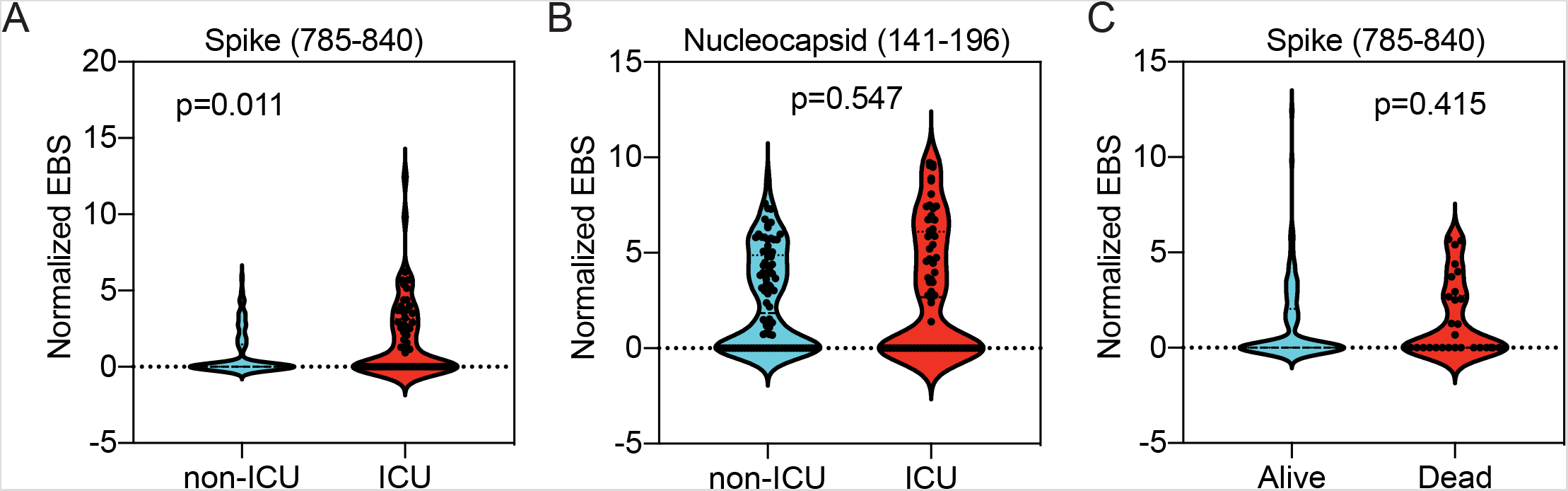
Prevalence of serological responses to SARS-CoV virus in hospitalized patients with ICU (A-B) or death status (C). The width of violin plots indicates the kernel density of values.

**Figure S3.**
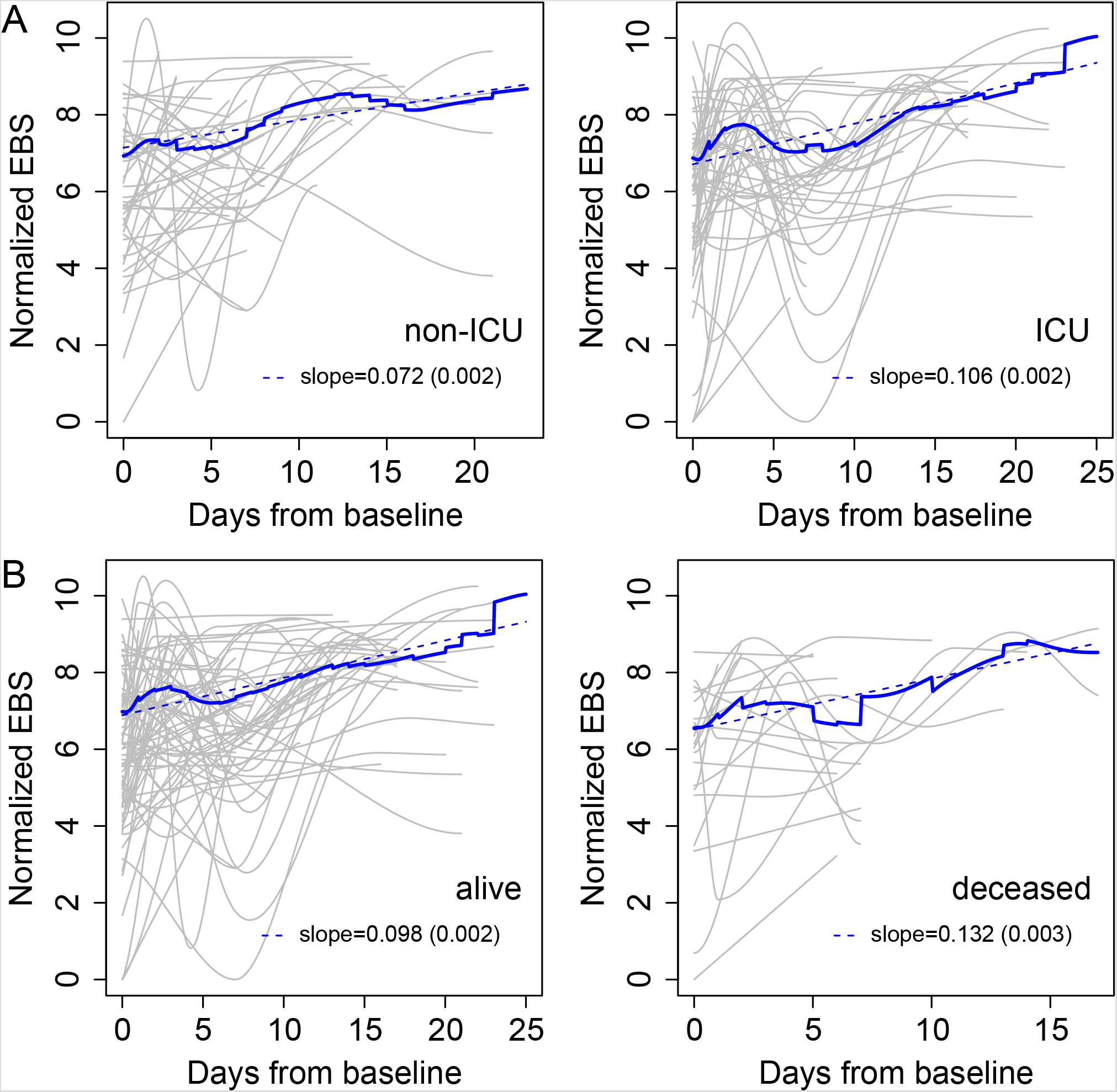
Longitudinal progression of the normalized EBS across individuals. (A) Individual trajectories over time for patients grouped by hospital ward (gray lines), which were averaged (solid blue line) and fitted by linear regression (dashed blue line; slope and standard error shown in the legend). Baseline refers to the first sample obtained after admission to the hospital. Left: non-ICU; right: ICU. (B) Analogous results for patients grouped by outcome. Left: alive; right: deceased.

**Figure S4.**
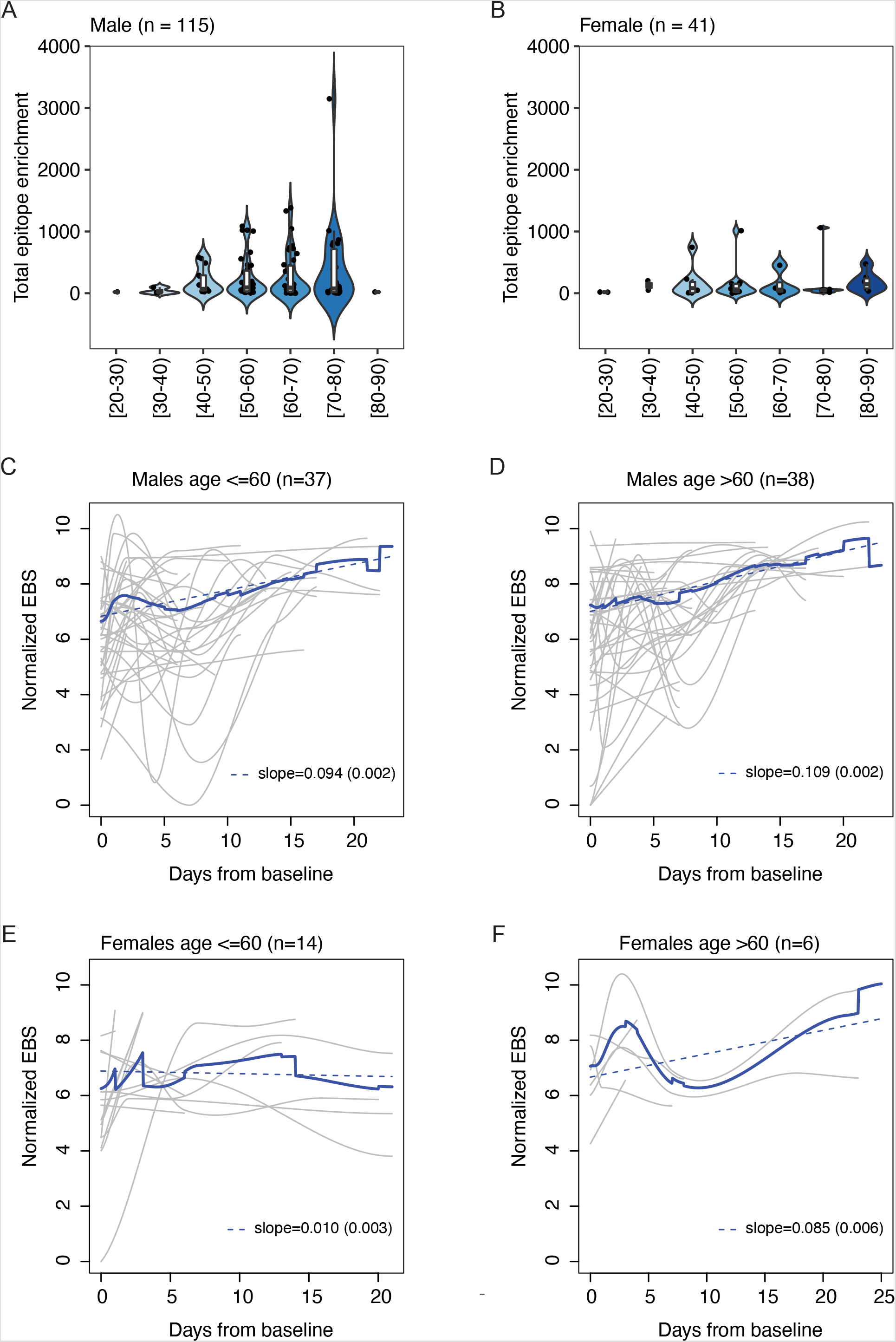
Sex and age effects of the humoral immune response of COVID-19 patients. (A-B) Total epitope enrichment at baseline as a function of age for male (A) and female(B) patients, respectively. (C-F) Longitudinal progression of the normalized EBS for younger males (C), older males (D), younger females (E), and older females (F), respectively. In violin plots, boxes span the interquartile range; lines within boxes represent the median; the width of violin plots indicates the kernel density of values.

**Table S1.** Viral prevalence in Brescia cohort. Related to Figure 1.

| **Virus species** | **Total**<br>**(n=156)** | **Moderate**<br>**(n=11)** | **Severe**<br>**(n=19)** | **Critical**<br>**(n=126)** |
| --- | --- | --- | --- | --- |
| Human respiratory syncytial virus | 81% | 64% | 79% | 83% |
| Human herpesvirus 4 | 74% | 91% | 95% | 70% |
| Human herpesvirus 1 | 67% | 55% | 79% | 66% |
| Human herpesvirus 5 | 55% | 55% | 63% | 54% |
| SARS-CoV | 48% | 27% | 53% | 49% |
| Rhinovirus B | 37% | 27% | 58% | 34% |
| Rhinovirus A | 33% | 27% | 47% | 32% |
| Human immunodeficiency virus 1 | 23% | 27% | 32% | 21% |
| Human herpesvirus 3 | 22% | 9% | 32% | 22% |
| Influenza B virus | 19% | 9% | 16% | 21% |
| Human adenovirus C | 17% | 9% | 21% | 17% |
| Enterovirus B | 14% | 0% | 16% | 15% |
| Enterovirus C | 13% | 0% | 16% | 14% |
| Dengue virus | 13% | 9% | 21% | 12% |
| Human herpesvirus 6B | 13% | 9% | 11% | 13% |
| Mamastrovirus 1 | 10% | 9% | 11% | 10% |
| Orf virus | 10% | 0% | 16% | 10% |
| Betacoronavirus 1 | 10% | 0% | 11% | 10% |
| Rotavirus A | 10% | 0% | 11% | 10% |
| Enterovirus A | 8% | 0% | 16% | 7% |
| Influenza A virus | 8% | 9% | 0% | 9% |
| Human adenovirus D | 7% | 18% | 0% | 7% |
| Human coronavirus NL63 | 7% | 0% | 11% | 7% |
| Human herpesvirus 8 | 6% | 9% | 0% | 7% |
| Norwalk virus | 6% | 0% | 21% | 5% |
| Hepatitis B virus | 6% | 0% | 11% | 6% |
| Hepatitis C virus | 6% | 0% | 0% | 7% |
| Human herpesvirus 2 | 6% | 9% | 0% | 6% |
| Human coronavirus 229E | 4% | 0% | 5% | 4% |
| Alphapapillomavirus 9 | 3% | 0% | 5% | 3% |

